# Identifying Blood Proteomic Markers of Parkinson’s Disease Dementia Using High-Throughput Approaches

**DOI:** 10.64898/2026.06.30.26356774

**Authors:** Raquel Real, Rafaela Ravazio, Anahita Nodehi, Yoav Ben-Shlomo, Nigel Williams, Rodrigo C. Barros, Donald Grosset, Michele Hu, Laura Winchester, Huw R. Morris

## Abstract

**INTRODUCTION:** Parkinson’s disease (PD) presents with motor and non-motor symptoms, including dementia, but the severity and rate of cognitive decline are heterogeneous and difficult to predict clinically.

**METHODS:** We quantified baseline serum proteins with the high-throughput SomaScan^®^ assay in 834 PD individuals and performed Cox regression to identify proteins associated with subsequent development of dementia. Candidate biomarker proteins were replicated in 371 individuals from an independent cohort and meta-analysed.

**RESULTS:** Protein targets significantly associated with progression to dementia were predominantly involved in synaptic plasticity, protein degradation/lysosomal function and extracellular matrix organisation. Mendelian Randomisation further revealed that changes in the Nogo receptor RTN4R may be causally associated with the development of Lewy body dementia.

**DISCUSSION:** We identified several proteins predicting progression to dementia in PD, indicating changes in blood proteome that precede the development of clinical symptoms by several years, providing a window of opportunity to identify at-risk individuals early on.

## 1. Background

Parkinson’s disease (PD) is a common neurodegenerative disorder, predicted to affect over 25 million people worldwide by 2050, due in part to population aging.^1^ Although PD is typically characterised by motor symptoms, non-motor symptoms such as cognitive impairment and dementia are common. The rate of dementia varies across studies, with risk estimated to be 50% after 10-15 years of motor symptom onset.^2^ Several clinical risk factors for the development of dementia have been described and include older age and age at PD onset, longer duration of PD, worse disease severity, non-tremor subtypes, the presence of REM sleep behaviour disorder, orthostatic hypotension and neuropsychiatric manifestations including hallucinations.^3,4^ Genetic risk factors have also been identified that increase the risk of dementia, the most consistent being the *APOE* ε4 allele.^5–8^ Risk variants in several other genes have been identified in single studies.^8,9^

The Movement Disorders Society has established clinical diagnostic criteria for dementia associated with PD, which is characterised by predominant impairment in attention, executive and visuospatial functions.^10^ Free recall memory impairment which improves with cueing can also be present, along with neurobehavioral features, including apathy, depressed or anxious mood, hallucinations and delusions.

Recognising which individuals with PD are at increased risk of developing dementia is a significant challenge, particularly early in the disease course. In the field of Alzheimer’s disease (AD), several CSF and more recently blood-based protein biomarkers have been identified that can accurately predict conversion from cognitively normal and mild cognitive impairment to dementia.^11,12^ Small targeted studies have suggested that AD-related plasma biomarkers and plasma neurofilament light chain (NfL) may predict cognitive decline in PD.^13,14^ However, no clinically validated biomarkers currently exist for predicting Parkinson’s disease dementia. Here, we aim to identify proteomic profiles associated with development of dementia in recently diagnosed PD patients by using high throughput proteomic assays. This strategy has the potential to identify protein markers that could be developed into clinical assays for risk prediction and diagnosis, as well as highlighting disease mechanisms underlying the development of dementia in PD and potential future treatment strategies.

## 2. Methods

### 2.1. Study Participants

The discovery cohort consisted of the Tracking Parkinson’s study (TPD), a UK-based multicentre, observational, longitudinal study, active between 2012 and 2022.^15^ Parkinson’s disease patients with recent symptom onset (<3.5 years) or young onset (age <50 years old) were recruited across 72 specialist clinics. Blood samples were collected at baseline and participants assessed with validated clinical scales every 18 months.

The replication cohort consisted of the Oxford Parkinson’s Disease Centre Discovery (OPDC) cohort,^16^ a multicentre UK-based observation, longitudinal study. Idiopathic Parkinson’s disease patients who were diagnosed within the last three years were recruited from 11 participating centres. Clinical assessments were performed at 18-month intervals. Both studies were carried out in accordance with the Declaration of Helsinki and were approved by the respective Multicentre Research Ethics Committee. All patients provided written informed consent.

In all cohorts, cognitive status at each study visit was defined based on the Movement Disorders Society (MDS) Task Force recommendations^17^ using previously described criteria.^8^ Specifically, we classified participants as having PD dementia if they had (1) adjusted MoCA scores ≤21/30; (2) impairment in at least two cognitive domains (attention/serial sevens ≤2/3; language/letter fluency 0/1; memory/delayed recall ≤4/5; visuospatial/executive ≤4/5); (3) and a cognitive deficit severe enough to impact activities of daily living (MDS-Unified Parkinson’s disease Rating Scale (UPDRS) 1.1 score ≥2), in the absence of severe depression (MDS-UPDRS 1.3 score <4). In addition, participants who withdrew from the study due to dementia were also classified as having a diagnosis of PD dementia. Individuals with missing data that meant that cognitive status could not be ascertained were excluded from subsequent analyses.

### 2.2. *APOE* genotyping

DNA extracted from whole blood was genotyped in the TPD cohort using the Illumina HumanCoreExome array and in the OPDC cohort using either the Illumina HumanCoreExome-12 v1.1 or the Illumina Infinium HumanCoreExome-24 v1.1 arrays. After standard quality control^8^ genotypes were imputed using the Michigan Imputation Server (https://imputationserver.sph.umich.edu) against the Haplotype Reference Consortium reference panel (v.r1.1 2016; http://www.haplotype-reference-consortium.org/). *APOE* genotypes were inferred from the imputed genotypes of the rs7412 and rs429358 variants. Participants were classified as *APOE* ε4 carriers if they carried at least one ε4 allele.

### 2.3. Proteomics assay

We measured protein levels in serum samples from 846 participants of the TPD cohort and from 373 participants of the OPDC cohort at baseline. Briefly, 10 mL of venous blood was collected in SST tubes and centrifuged at 2500g for 15 min, on average within 1 hour of collection. Serum aliquots were then stored in cryotubes at-80°C until further processing. Protein levels were measured using the SomaScan^®^ multiplex assay (v3.2, SomaLogic^®^, Inc., Boulder, CO).^18^ After processing using SomaLogic’s proprietary normalisation pipeline, protein concentration measurements were obtained in Relative Fluorescent Units (RFU). We used the R package *SomaDataIO* (v6.4.0) to read in and process the SomaScan^®^ data. RFU values were log2-transformed. Non-human analytes were excluded from the analysis (*n* = 8). Samples that did not meet a sample normalization scale factor check were excluded from the analysis (*n* = 3 from the TPD cohort and *n* = 2 from the OPDC cohort). Outlier samples were identified based on mean protein levels ± 3*interquartile range (IQR) and also excluded from downstream analyses (*n*= 4 in the TPD cohort; Supplementary Figure 1). After merging with the clinical data, 834 participants with PD from the TPD cohort and a further 371 participants from the OPDC cohort were available for analysis. A total of 3,998 aptamers were analysed.

### 2.4. Statistical Analyses

All statistical analyses were conducted in RStudio (version 2025.9.0.387). *P*-values were adjusted for multiple testing using the Benjamini–Hochberg (BH) method. *P*-values <0.05 were considered nominally significant, with FDR-adjusted *P* < 0.05 considered statistically significant. We used Pearson’s Chi-squared tests or Wilcoxon rank sum tests to compare categorical or continuous clinical variables between groups, respectively.

#### 2.4.1 Survival analysis

We used Cox proportional hazards models to perform survival analysis. Time-to-dementia was calculated as the number of years from PD diagnosis to the first visit when dementia criteria were met, or to study withdrawal due to dementia. Cases who met dementia criteria at baseline were excluded from the survival analysis (*n* = 24 in the discovery cohort and *n* = 2 in the replication cohort). To identify proteins associated with dementia-free survival, we first used the *coxph* function from the *survival* package (v3.8.3) in the TPD cohort. Time-to-dementia was regressed against the levels of each analyte separately, with sex and age at baseline used as covariates. We used the *sva* package (v3.57.0) to estimate surrogate variables (SVs) as latent variables that might remove potential batch effects or other sources of unwanted variation; the first three SVs were also included as covariates in our models. Nominally significant analytes from the discovery phase (*n* = 335) were then replicated in the OPDC cohort using the same covariates. Finally, results from the discovery and replication cohorts were meta-analysed using the *metafor* package (v4.8-0). Heterogeneity across studies was assessed using Cochran’s Q-test, I² and Tau². A sensitivity analysis using *APOE* ε4 status as a covariate was also performed in both cohorts and then meta-analysed.

#### 2.4.2 Differential abundance analysis stratified by *APOE* genotype status

We used the *limma* package (v3.65.4) to perform a differential expression analysis between *APOE* ε4 carriers and non-carriers in the TPD cohort, using sex and age at baseline as covariates. Nominally significant analytes (*n* = 218) were then analysed in the OPDC cohort and results meta-analysed with those from the discovery cohort.

#### 2.4.3 Mediation analysis

We performed causal mediation analysis using the *cmest* function from the *CMAverse* package (v0.1.0) to evaluate whether baseline protein abundance mediated the association between *APOE* ε4 status and dementia risk in the discovery cohort (TPD). A regression-based mediation framework was applied, modelling the mediator using linear regression and the outcome using a Cox proportional hazards model. Models were adjusted for sex and age at baseline. Protein abundance values were standardised prior to analysis, and participants with missing data were excluded. Standard errors and confidence intervals of causal effects were estimated using bootstrap inference (1000 iterations).

#### 2.4.4. pQTL analysis

To test the pQTL between the *APOE* ε4-tagging SNP rs429358 and serum levels of selected proteins in the discovery cohort (TPD), we obtained genetic principal components (PCs) in *plink* v1.9 (https://www.cog-genomics.org/plink/) and performed linear regression adjusted for sex, age at baseline and the first two PCs.

#### 2.4.5 Pathway enrichment and protein clustering analysis

We used the *clusterProfiler* package (v4.18.4) to make Gene Ontology (GO) and REACTOME functional annotations of the proteins identified in the APOE-adjusted survival meta-analysis. Redundant terms were removed using the *simplify* function. *STRINGdb* package (v2.21.0) was then used to obtain protein clusters using the *fastgreedy* algorithm. Protein clusters were also functionally annotated with GO on biological processes (GO:BP).

#### 2.4.6 Two-Sample Mendelian Randomisation

We ran two-sample *cis*-MR analysis with the *TwoSampleMR* package (v0.6.22), using protein quantitative trait loci (pQTL) obtained from Hawkes *et al* as genetic instruments and GWAS summary statistics from dementia traits as the outcome.^8,19,20^ For each protein, we selected genetic instruments within a ±1 Mb *cis* window, filtering for SNPs with genome-wide significance (*P* < 5×10^-08^) and *F*-statistic ≥ 10. Independent SNPs were then selected by performing LD clumping with default settings (r² < 0.001, ± 10 Mb window), using the 1000 Genomes European reference panel. We harmonised the datasets using the *harmonise_data* function and excluded palindromic variants. Causal effects were estimated by random-effects inverse-variance weighted (IVW) MR when more than one suitable genetic instrument was available, otherwise the Wald ratio was used. Sensitivity analyses were performed using weighted median and/or weighted mode when appropriate. Directionality was assessed using Steiger test, horizontal pleiotropy was evaluated with the MR-Egger intercept test and heterogeneity was assessed with Cochran’s Q test, depending on instrument counts.

### 2.5. Data availability

Proteomics data is available on the AD Discovery Portal (https://discover.alzheimersdata.org) via application to the Global Neurodegeneration Proteomics Consortium (GNPC, https://www.neuroproteome.org/).^21^

Clinical data from TPD and OPDC cohorts is available through the CPP Integrated Parkinson’s Database (https://c-path.org/tools-platforms/integrated-parkisons-database/). Code used in preparation of this manuscript is available on GitHub (https://github.com/raquelreal/PDD_SomaScan_Proteomics) and given a persistent identifier (doi:10.5281/zenodo.21004766). A Key Resources Table can be found in supplementary material (Supplementary Table 1).

## 3. Results

### 3.1. Cohort characteristics

An overview of the study design is shown in Figure 1. Patient demographics and clinical features are described in Table 1. In both discovery and replication cohorts, participants who developed dementia were older at disease onset and at the baseline assessment. The dementia groups had the highest proportion of male participants and of *APOE* ε4 allele carriers, consistent with previous literature.^5,7,22–24^ The median (IQR) time between baseline serum sampling and the development of dementia was 3.11 (2.97) and 3.79 (3.80) years in the TPD and OPDC cohorts, respectively.

**Figure 1.**
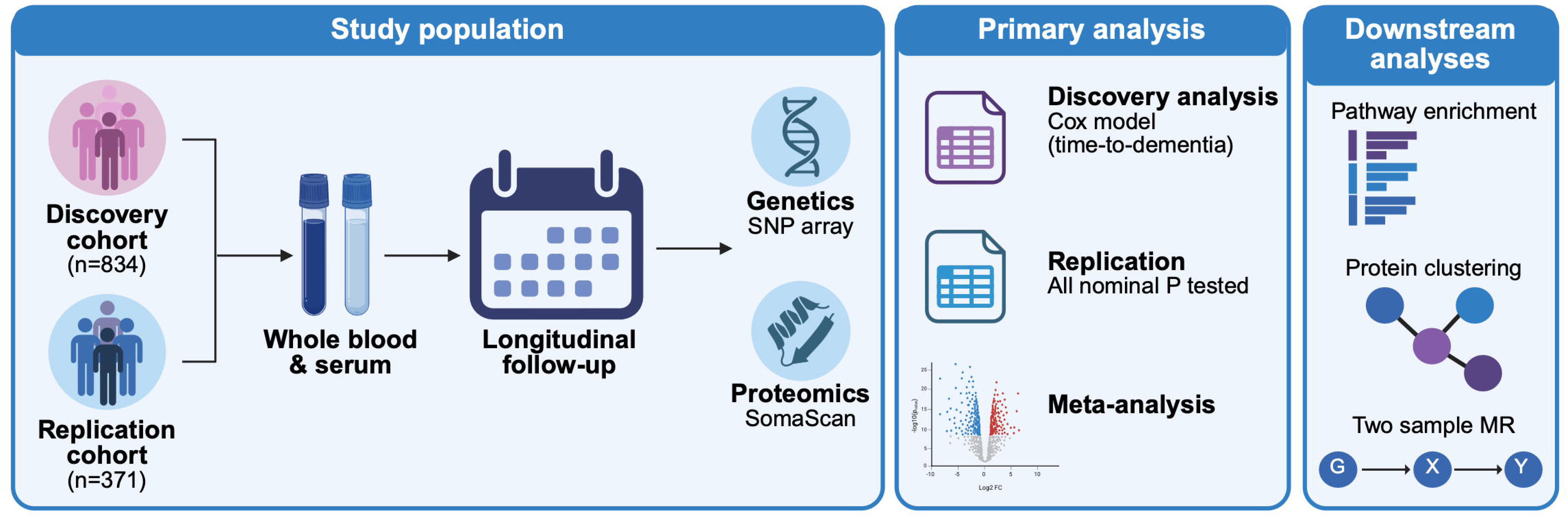
Study Design, including the study population and overview of the statistical analyses performed. Created in BioRender https://BioRender.com/3uni34f.

**Table 1.** Demographic and clinical features of study participants.

|  |  | Discovery |  |  |  | Replication |  |  |  |
| --- | --- | --- | --- | --- | --- | --- | --- | --- | --- |
|  |  | PD total<br>N=834 | PD<br>N=678 | PDD<br>N=83 | P-value | PD total<br>N=371 | PD<br>N=303 | PDD<br>N=58 | P-value |
| Sex, n (%) | Female | 308 (36.9) | 272 (40.1) | 18 (21.7) | 0.001 | 137 (36.9) | 119 (39.3) | 15 (25.9) | 0.053 |
|  | Male | 526 (63.1) | 406 (59.9) | 65 (78.3) |  | 234 (63.1) | 184 (60.7) | 43 (74.1) |  |
| Age onset, median (IQR) |  | 65.2 (12.3) | 64.4 (12.3) | 68.6 (8.7) | 3.73e-06 | 65.4 (12.9) | 64.9 (14.1) | 69.4 (11.6) | 1.76e-04 |
| Age baseline, median (IQR) |  | 68.1 (11.9) | 67.0 (11.9) | 72.1 (7.8) | 3.66e-08 | 68.1 (13.3) | 67.5 (3.6) | 72.3 (11.7) | 1.56e-04 |
| Disease duration baseline, median (IQR) |  | 1.1 (1.6) | 1.1 (1.6) | 1.1 (1.5) | 0.53 | 0.70 (0.9) | 0.73 (1.0) | 0.65 (0.9) | 0.84 |
| MoCA baseline, median (IQR) |  | 26 (5) | 26 (4) | 22 (4) | 2.80e-13 | 26 (5) | 26 (4) | 23 (4) | 2.72e-08 |
| APOE status,<br>n (%) | ε4 carrier | 217 (26.0) | 167 (24.6) | 35 (42.2) | 1.94e-04 | 88 (23.7) | 60 (19.8) | 24 (41.4) | 2.63e-04 |
|  | ε4 non-carrier | 525 (62.9) | 452 (66.7) | 38 (45.8) |  | 246 (66.3) | 213 (70.3) | 28 (48.3) |  |
|  | Missing | 92 (11.0) | 59 (8.7) | 10 (12.0) |  | 37 (10.0) | 30 (9.9) | 6 (10.3) |  |

### 3.2. Survival analysis of time-to-dementia

To identify proteins associated with progression to dementia, we first fitted Cox proportional hazards models in the discovery cohort, which identified 335 nominally significant protein targets (Figure 2A; Supplementary Table 2). These targets were subsequently used to fit Cox models in the replication cohort (Supplementary Table 3). The meta-analysis of results from the two cohorts identified 32 statistically significant protein targets after correction for multiple testing (Table 2; Supplementary Table 4). All the protein targets that were associated with progression to dementia in the meta-analysis had the same direction of effect in the discovery and replication cohorts (Figure 2B). In addition, there was strong correlation between the z-score of the effect sizes of significant proteins in both cohorts (Pearson’s correlation: *r* = 0.90, *P* = 3.69e-12).

**Figure 2.**
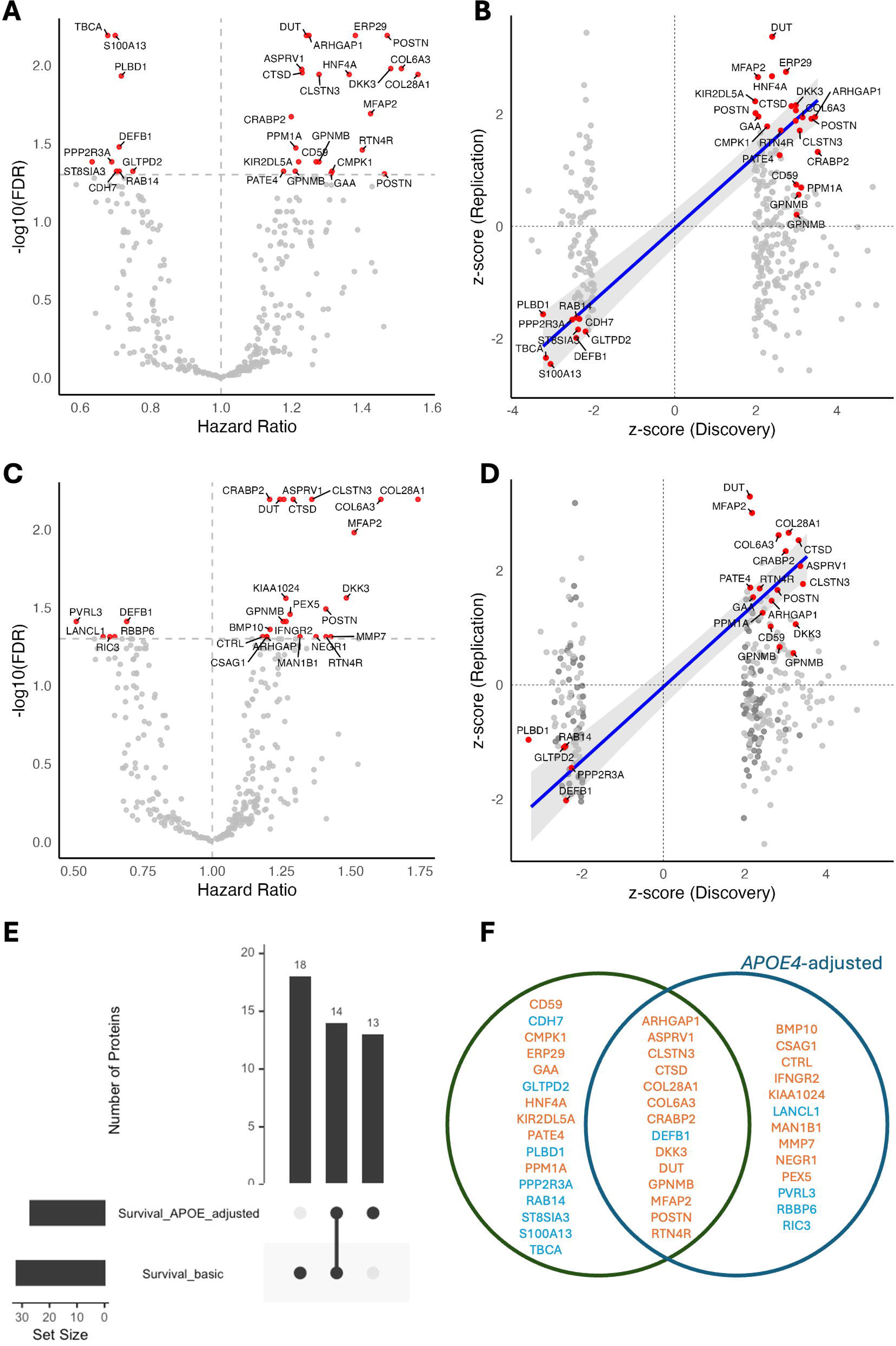
Survival analysis of progression to dementia in PD. **(A-C)** Volcano plot of meta-analysis without (A) and with (C) *APOE* ε4 as a covariate. Colour scheme: red, FDR-adjusted *P* < 0.05; grey, FDR-adjusted *P* > 0.05. **(B-D)** Correspondence plot between the z-score of the estimate in the Discovery and Replication cohorts without (B) and with (D) *APOE* ε4 as a covariate. Colour scheme: red, FDR-adjusted *P* < 0.05; grey, FDR-adjusted *P* > 0.05. **(E)** Upset plot of meta-analysis with and without *APOE* ε4 as a covariate. **(F)** Venn diagram of FDR-significant proteins identified in the survival meta-analysis with (right) and without (left) *APOE* ε4 as a covariate. Proteins with increased abundance are represented in orange and proteins with decreased abundance are represented in blue.

**Table 2.**
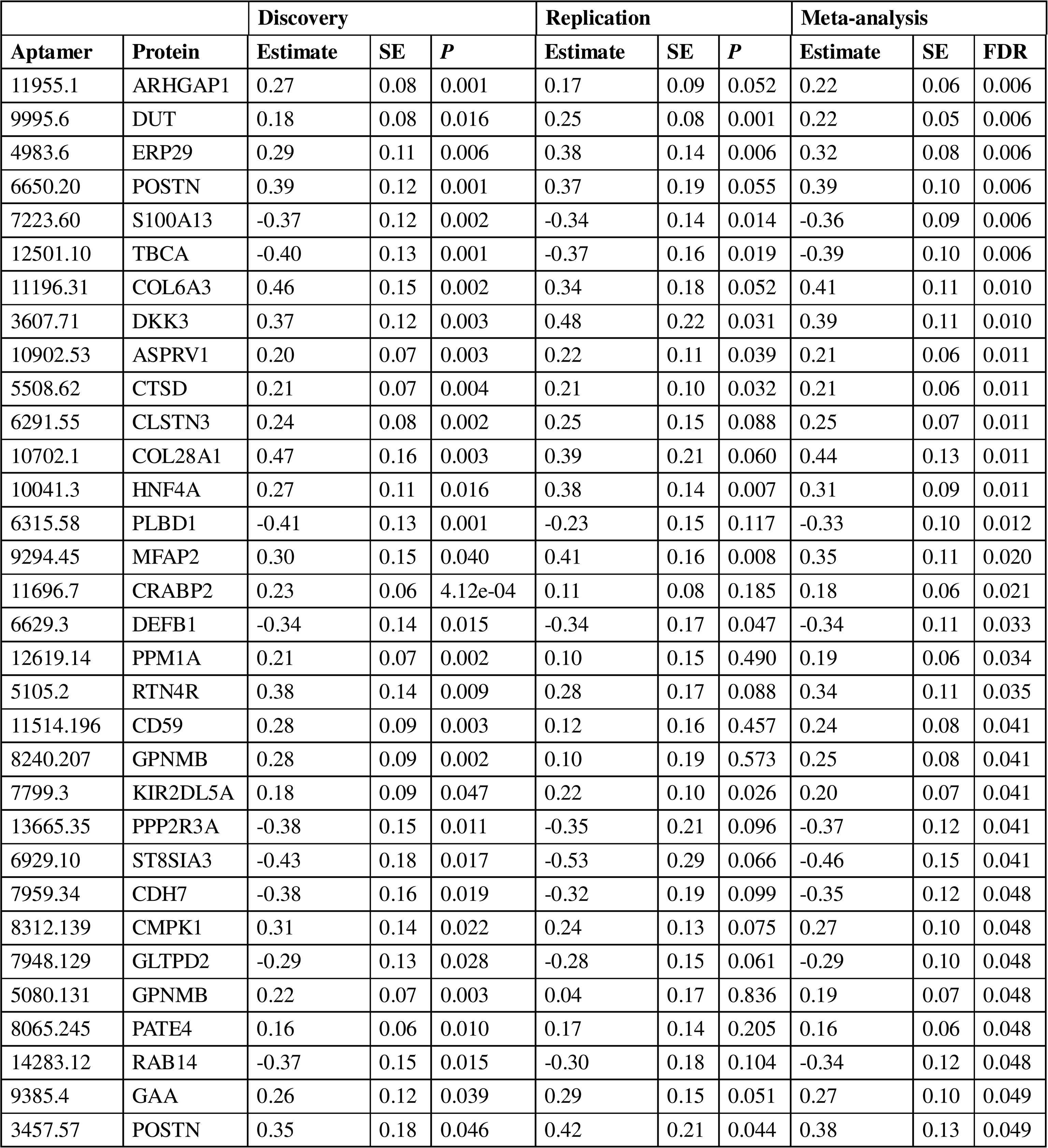
Significant proteins from CPH time-to-dementia meta-analysis.

### 3.3. Differential protein abundance stratified by *APOE* genotype status

Given that the *APOE* ε4 genotype was more common in PD individuals who developed dementia (Table 1) and has previously been associated with the risk of conversion to dementia in PD,^6–8^ we explored differential protein abundance in PD individuals with and without the *APOE* ε4 allele (Figure 3, Supplementary Tables 5-7). Interestingly, two of the proteins identified in the survival meta-analysis (TBCA and S100A13) were also differentially abundant in *APOE* ε4 allele carriers. TBCA and S100A13 levels showed a strong negative correlation with the *APOE* ε4-tagging SNP (rs429358) in both cohorts (TBCA: *r* =-0.61, *P* = 2.77e-73 (TPD); *r* =-0.57, *P* = 1.31e-29 (OPDC); S100A13: *r* = - 0.61, *P* = 7.73e-75 (TPD); *r* =-0.54, *P* = 1.52e-26 (OPDC)). In addition, there was a dose-dependent association of rs429358 with protein levels of TBCA and S100A13 (Supplementary Figure 2A), reflecting known trans pQTL effects of the *APOE* locus on these proteins in both plasma and CSF.^25,26^ To test whether the effect of *APOE* ε4 on dementia risk was mediated by circulating levels of these proteins, we performed mediation analysis, which provided no evidence that TBCA or S100A13 significantly mediate the effect of *APOE* ε4 on dementia risk (Supplementary Table 8). None of the other proteins differentially abundant in *APOE* ε4 carriers significantly associate with dementia outcomes in PD patients (Supplementary Figure 2B).

**Figure 3.**
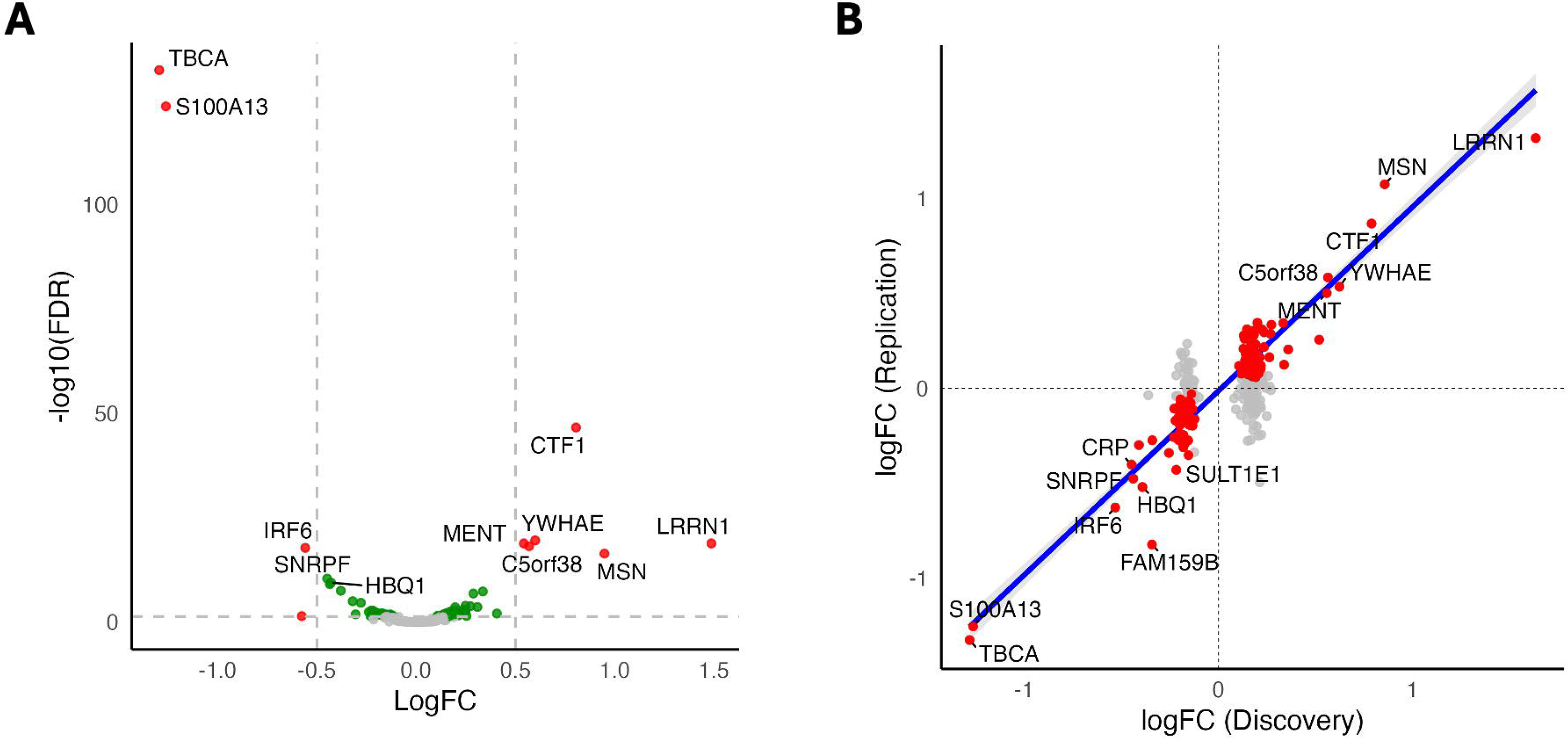
Differential abundance analysis in PD patients stratified by APOE4 status. **(A)** Volcano plot of meta-analysis. Colour scheme: red, FDR-adjusted *P* < 0.05 and |logFC| > 0.5; green, FDR-adjusted *P* < 0.05 and |logFC| < 0.5; grey, FDR-adjusted *P* > 0.05. **(B)** Correspondence plot between the logFC in the Discovery and Replication cohorts. Colour scheme: red, FDR-adjusted *P* < 0.05; grey, FDR-adjusted *P* > 0.05.

Given that serum changes to TBCA and S100A13 levels in individuals with PD who developed dementia is confounded by their *APOE* ε4 status, we performed a sensitivity survival analysis using the *APOE* genotype as an additional covariate. As expected, the association of TBCA and S100A13 levels with dementia outcomes was attenuated after adjusting for *APOE* genotypes (Figure 2C-D). However, fourteen of the original protein targets remained significant and 13 additional targets were identified in the meta-analysis of the discovery and replication cohorts after correcting for *APOE* ε4 status (Figure 2E-F; Supplementary Tables 9-11).

### 3.4 Pathway enrichment analysis

Next, we performed pathway enrichment analysis using FDR-significant protein targets from the survival meta-analysis adjusted for *APOE* genotype, which demonstrated enrichment in pathways involved in extracellular matrix organisation and regulation of synapse organisation (Figure 4A). Clustering using a STRING network of protein-protein interactions revealed that proteins cluster into three groups (Figure 4B). Functional annotation using Gene Ontology showed that cluster 1 was comprised of proteins involved in extracellular matrix organisation, while cluster 2 had proteins linked to immune responses in the context of lysosomal stress and cluster 3 proteins were involved in neural processes, such as regulation of synapse assembly, synaptic transmission and neuron differentiation (Supplementary Table 12).

**Figure 4.**
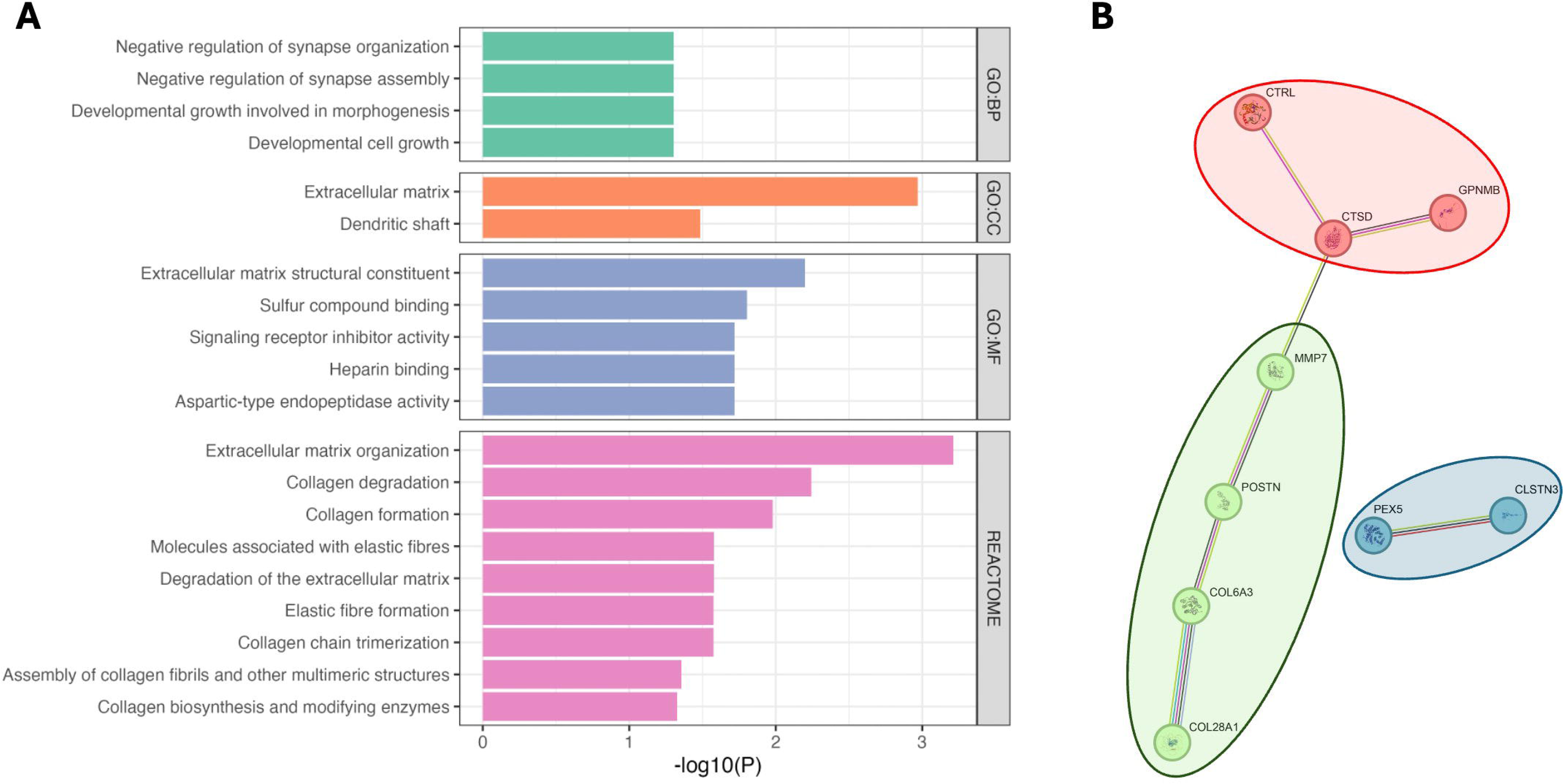
Pathway enrichment analysis of proteins associated with dementia-free survival. **(A)** Enriched GO and Reactome pathways with FDR-adjusted *P* < 0.05 and gene count > 1. **(B)** String-db network and clustering using fastgreedy. Colours indicate the three identified clusters. Cluster 1 (green) contains proteins linked to extracellular matrix organization (cell-matrix adhesion, collagen metabolic process), cluster 2 (red) is composed of proteins involved regulation of immune responses (antigen presentation, T cell proliferation, tumour necrosis factor production, mononuclear cell proliferation), and cluster 3 (blue) has proteins linked to regulation of neural processes (synapse assembly, synaptic transmission, neuron differentiation, neuron projection extension).

### 3.5 Mendelian Randomisation

To investigate potential causal relationships between proteins identified in the survival meta-analysis and the development of related dementia traits, we performed two-sample *cis*-MR analysis. Suitable genetic instruments (pQTLs) were identified for 15 proteins (Supplementary Table 12). We found evidence of a causal association between genetically predicted higher levels of RTN4R and an increased hazard of developing dementia with Lewy bodies (OR [95% confidence interval] = 1.35 [1.10-1.65]; adjusted-*P* = 0.042). This association was robust to potentially pleiotropic instruments, as demonstrated by consistent effects using the weighted median method (*P* = 0.030).

Sensitivity analyses showed no evidence of heterogeneity (Cochran’s Q test; Supplementary Table 14) or directional pleiotropy (MR-Egger intercept test; Supplementary Table 15). Leave-one-out analysis confirmed that no single SNP disproportionately influenced the estimate (Supplementary Table 16). Additionally, the Steiger test confirmed the validity of the instruments for the exposure (Supplementary Table 17). No significant associations were found between genetically predicted protein levels and the risk of PD dementia or Alzheimer’s disease after correction for multiple testing.

## 4. Discussion

Dementia is a frequent complication in PD patients, but the timing and rate of cognitive decline vary widely between individuals. Our understanding of this heterogeneity is limited, and we currently lack the tools to identify high-risk individuals early in the disease course. Overall, cognitive symptoms in PD correlate with the neocortical burden of Lewy bodies;^27,28^ however, Alzheimer’s type pathology is also a frequent finding in the brains of patients with PD dementia and is thought to contribute to the cognitive symptoms.^29^ In fact, co-pathology occurs in as many as 50% of cases with Parkinson’s disease dementia.^30^ Recently, α-synuclein seed amplification assays (SAA) in CSF have been developed to identify individuals with brain Lewy body pathology.^31^ Interestingly, it was shown that faster seeding kinetics predict cognitive decline in PD independently of AD co-pathology.^32^ But while blood-based biomarkers to detect AD-type amyloid pathology have been developed and are being increasingly used in clinical settings,^12^ similar biomarkers specifically linked to neocortical Lewy body pathology are still lacking. Here, we explored the proteomic profile of PD dementia in a large cohort to identify protein changes associated with the development of dementia, which could provide insights into the pathophysiology of dementia in PD, as well as identify potentially novel blood-based biomarkers.

We have identified several dysregulated proteins in PD individuals who later went on to develop dementia, which suggests that blood proteomic changes precede the development of dementia by several years. Overall, dysregulated proteins fall into one of the following categories: (1) synaptic plasticity (e.g., RTN4R, NEGR1, CLCTN3); (2) lysosomal dysfunction (e.g., GPNMB, CTSD); and (3) extracellular matrix constituents and regulators (e.g., COL28A1, COL6A3, MMP7, MFAP2, POSTN).

Synapse loss and changes in synaptic plasticity are hallmarks of dementia. RTN4R (also known as the Nogo receptor, or NgR) is highly expressed in neurons, where it regulates the development of synaptic circuits by regulating dendritic arborisation, axonal elongation and synapse formation.^33^ Increased expression of RTN4R has been shown in the hippocampus of AD patients, where it colocalised with neurofibrillary tangles.^34^ In addition, higher CSF RTN4A levels, a ligand of RTN4R, have been detected in patients with AD and PD compared to controls.^35^ We have identified increased serum levels of RTN4R in our survival analysis, with MR analysis further suggesting a causal relationship between genetically determined higher levels of RTN4R and increased risk of Lewy body dementia, which encompasses both PD dementia and dementia with Lewy bodies (DLB). The exact mechanism by which RTN4R levels increase the risk of dementia are not fully elucidated, but previous studies have demonstrated that Nogo signalling via RTN4R negatively impacts structural synaptic plasticity and dendritic complexity, restricts excitatory synapse formation, and that overexpression of RTN4R impairs the formation of long-term memories.^36–38^

*GPNMB* has been consistently identified in case-control PD GWAS studies as a risk factor for PD.^39–41^ It is secreted by microglia in response to lysosomal stress,^42^ and levels of GPNMB have been shown to be increased in AD and other neurodegenerative conditions.^43^ The role of GPNMB in PD pathophysiology is still under debate. It has been suggested that secreted GPNMB could have a neuroprotective role by reducing astrocyte-mediated neuroinflammation,^44^ while a separate study found that GPNMB contributes to the clearance of oligomeric amyloid β_1-42_ peptides by microglia.^45^ Contrary to these putative neuroprotective roles, GPNMB has also been found to interact with α-synuclein, which was necessary for fibrillar α-synuclein internalization and development of α-synuclein pathology.^46^

CTSD is a lysosomal protease involved in the degradation of several protein substrates, including α-synuclein.^47,48^ Therefore, the association of increased serum levels of CTSD with an increased hazard of developing dementia in PD patients could reflect a compensatory mechanism to increase protein clearance, given that dementia closely relates to the extent of neocortical Lewy bodies.^27,28^ Interestingly, *CTSD* has also been identified in a genetic burden analysis of lysosomal genes in association with PD risk.^49^

In the central nervous system, the extracellular matrix acts as a scaffolding for synapse formation and plasticity, as well as regulating cell signaling in the brain and maintaining the integrity of the blood-brain barrier.^50^ Several proteins involved in extracellular matrix organization and remodeling have been identified in our analysis. Collagen VI has been shown to have a neuroprotective role against amyloid β toxicity and during physiological aging.^51,52^ Increased levels of COL6A3, a type of collagen VI chain, have also been found in the brains of PD patients, and it is possible that this similarly reflects a neuroprotective mechanism in PD.^53^

This study has several limitations. First, although we employed strict criteria for the classification of dementia cases based on the MDS Task Force recommendations, a clinical diagnosis of dementia based on in-person assessments was not available, with the potential for imprecise classifications. Second, our proteomic data is based on a single platform, and findings have not been orthogonally validated, although we have been able to use MR to generate genetic evidence for some of the targets. Third, the lack of data on amyloid co-pathology makes it impossible to discern the proteomic changes that are specific to Lewy body pathology. Additionally, although TPD and OPDC are broadly representative of the UK population, a further study of these findings in more diverse cohorts would complement the current results. Finally, our findings are based in serum and, despite indications from pathway enrichment results, might not reflect pathophysiological events in the brain.

In summary, we describe early blood proteome changes in a large longitudinal cohort of PD individuals at risk of PD dementia, including proteins involved in *APOE-*dependent and-independent pathways. Dysregulated proteins were mainly involved in synaptic plasticity, extracellular matrix structure and organisation, and in protein clearance mechanisms. Mendelian randomisation analysis suggested a causal association between Lewy body dementia risk and RTN4R, indicating that Nogo signalling might be an important pathophysiological mechanism in PD dementia. Our findings also suggest pathway overlap with other neurodegenerative disorders. For example, *NEGR1* is an Alzheimer’s disease risk locus,^54^ and it has also shown an association with the rate of cognitive decline in AD.^55^ In addition, we found an overlap with genetic loci previously identified in PD case-control studies (such as *GPNMB* and *CTSD*), suggesting that there might be some overlap between the risk of PD and PD dementia. It remains uncertain whether these dysregulated proteins are markers of disease state, reflecting early neurodegeneration in limbic and cortical areas that will eventually lead to dementia, or of disease pathogenesis, reflecting primary drivers of neurodegeneration. In some instances, protein changes likely reflect the upregulation of compensatory neuroprotective mechanisms. In any case, further research is necessary to investigate the value of these proteins as biomarkers for predicting risk of dementia early in the disease course.

## Acknowledgments

The authors would like to thank the patients for their time and samples donation. For the purpose of open access, the author has applied a CC BY public copyright licence to all Author Accepted Manuscripts arising from this submission.

## Conflict of interest statement

The authors have no conflicts of interest to report related to this work. Unrelated to this work, H.R.M. reports paid consultancy from Arvinas, Aprinoia, Skyhawk, AI Therapeutics, Neuron23; lecture fees/honoraria - Movement Disorders Society, Bial, Calico. H.R.M. is a co-applicant on a patent application related to C9orf72 - Method for diagnosing a neurodegenerative disease (PCT/GB2012/052140). Y.B.S. has received consultancy fees from Parkinson’s UK for epidemiological support.

## Funding sources

The Tracking Parkinson’s study was funded by Parkinson’s UK (J-1101). The Oxford Discovery Cohort is funded by Parkinson’s UK (J-2101- ‘Understanding Parkinson’s Progression’) and supported by the National Institute for Health Research (NIHR) Oxford Biomedical Research Centre based at Oxford University Hospitals NHS Trust and University of Oxford, and the NIHR Clinical Research Network: Thames Valley and South Midlands.

This research was funded in part by Aligning Science Across Parkinson’s [ASAP-000478] through the Michael J. Fox Foundation for Parkinson’s Research (MJFF) and supported by the Global Parkinson’s Genetics Program (GP2; https://gp2.org). GP2 is funded by the Aligning Science Across Parkinson’s (ASAP) (https://ror.org/03zj4c476) initiative and implemented by The Michael J. Fox Foundation for Parkinson’s Research (MJFF) (https://ror.org/03arq3225). For a complete list of GP2 members see https://doi.org/10.5281/zenodo.7904831.

R. Ravazio was financed by the Coordenação de Aperfeiçoamento de Pessoal de Nível Superior - Brasil (CAPES) - Finance Code 001.

A. Nodehi receives salary support from the Gatsby Charitable Foundation.

L. Winchester is funded by Alzheimer’s Research UK (ARUK-SRF2023B-007) and Michael

J. Fox Foundation (MJFF-022845).

Funding sources were not involved in the study design; collection, analysis and interpretation of data; writing of the report; nor the decision to submit the article for publication.

## Consent statement

All human subjects provided written informed consent.

## Notes

### Author Declarations

Tracking Parkinson's Disease has multi-centre research ethics approval from the West of Scotland Research Ethics Committee (REC reference: 11/AL/0163). Oxford Parkinson's Disease Centre has multi-centre research ethics approval from the South Central Oxford A Research Ethics Committee (REC reference: 16/SC/0108).

