## Supplementary Figure for "Identifying Blood Proteomic Markers of Parkinson’s Disease Dementia Using High-Throughput Approaches"

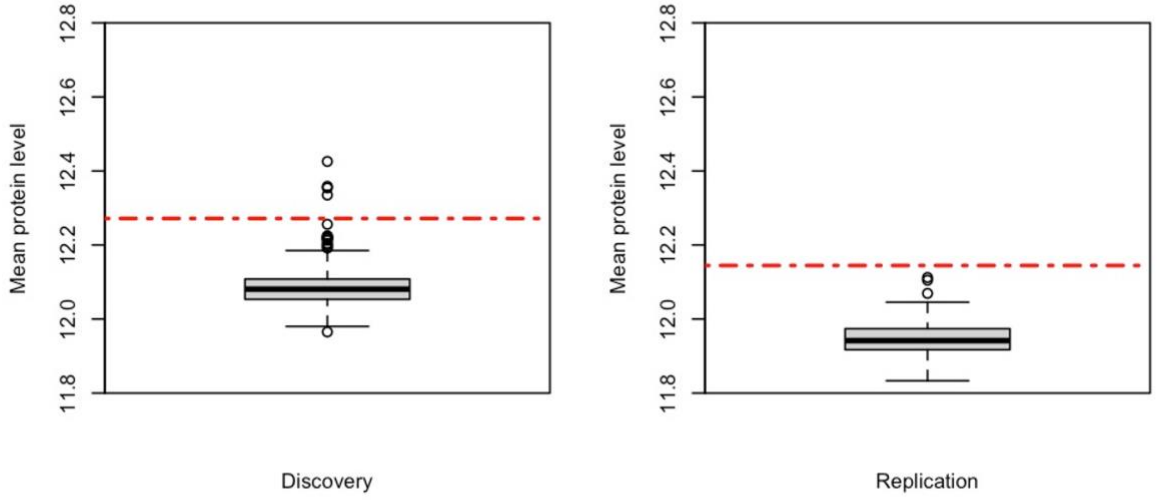


**Supplementary Figure 1.** Sample outliers in the Discovery (**left**) and Replication (**right**) cohorts. Outliers were identified as samples with mean protein levels outside the Q3 ± 3*IQR range.

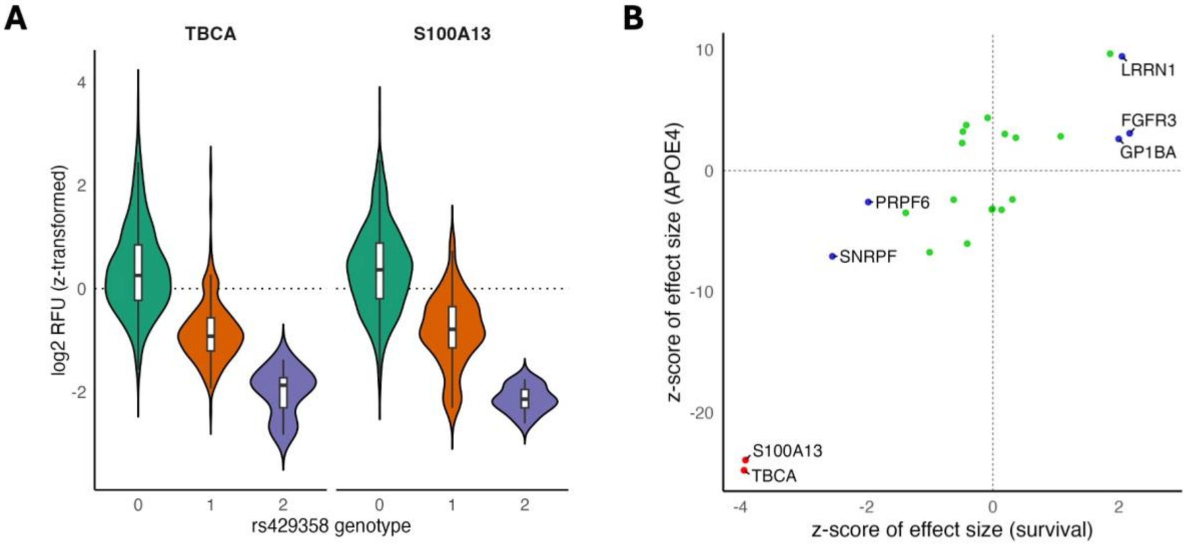


**Supplementary Figure 2. (A)** Violin plots of serum levels of TBCA/S100A13 by APOE4-tagging SNP rs429358 genotype. In a linear regression model adjusted for age at sampling, sex and the first two genetic PCs, rs429358 was strongly associated with levels of both proteins in the discovery cohort ($n$ = 718), with increasing copies of the alternative allele significantly associated with reduced levels of protein levels (TBCA: $\beta = -1.17$, se = 0.06, FDR-adjusted $P$ = 2.29e-73; S100A13: $\beta = -1.19$, se = 0.06, FDR-adjusted $P$ = 2.16e-74). **(B)** Correspondence plot between the z-score effect sizes of the survival meta-analysis and the APOE meta-analysis. Colour scheme: red, FDR-adjusted $P < 0.05$ in both meta-analyses; blue, FDR-adjusted $P < 0.05$ in the APOE meta-analysis and nominal $P < 0.05$ in the survival meta-analysis; green, FDR-adjusted $P < 0.05$ in the APOE meta-analysis and nominal $P > 0.05$ in the survival meta-analysis. There were no proteins in common that were significant only in the survival meta-analysis.
